# Role of personalized predictions for people with prediabetes: disseminating patient-centered estimates of benefit

**DOI:** 10.1101/2024.12.03.24318396

**Authors:** Natalia Olchanski, Elizabeth L. Ciemins, John Cuddeback, Francis Colangelo, Carolyn Koenig, David M. Kent

## Abstract

**Background:** To assess acceptability and feasibility of incorporating individualized risk prediction into clinical assessment, decision making, and communication of risk of type 2 diabetes and prevention recommendations.

**Methods:** We integrated a prediction model into the clinical workflow at a US health care organization. We conducted patient and provider focus groups and pre- and post-dissemination surveys and assessed the effect on referrals to and enrollment in the Diabetes Prevention Program, among 2,775 patients with prediabetes who had primary care visits between May 2018 and December 2020.

**Results:** Among patients with prediabetes seen in primary care during the study period, 79% had a calculation with the risk prediction model completed. After implementation of the risk prediction model, prevention intervention rates increased, with 62.3% of high-risk patients receiving an intervention within 1 year.

**Conclusions:** Used at the point of care during a shared decision-making discussion between the patient and provider, the diabetes risk calculator helped providers prioritize patients for diabetes prevention interventions, facilitated communication, and successfully improved rates of engagement in their care among patients with prediabetes.

## Background

Prediabetes is highly prevalent in the U.S., affecting approximately 98 million Americans.^1^ While persons with prediabetes are at an increased risk of developing type 2 diabetes, an individual’s risk depends on many factors and has been shown to vary widely. Our team has previously developed and validated clinical prediction models to estimate diabetes onset risk in patients identified as having prediabetes, as well as the risk-specific estimated treatment effect of an intensive lifestyle intervention, the Diabetes Prevention Program (DPP), and metformin, using both observational and randomized data.^2,3^ While it is not feasible to enroll everyone with prediabetes in the DPP, knowing individualized risk of diabetes and the potential benefits of preventive interventions might help health care providers prioritize patients to optimize the number of type 2 diabetes cases prevented when capacity is constrained.^4–6^ In this study we assessed the acceptability and feasibility of incorporating individualized risk prediction into clinical assessment, decision-making, and communication of type 2 diabetes risk and prevention recommendations. We also assessed the impact of implementing the risk model on referrals and enrollment in the DPP at one U.S. health care organization.

## Methods

### Study setting and population

We conducted this study at Premier Medical Associates (PMA), a 100-provider multi-specialty medical group in Pittsburgh, PA. The study sample included patients aged 18-75 with prediabetes, with no diagnosis of diabetes on their EHR problem list and their last HbA1c within a year prior to the visit, in the range 5.7-6.4% (39-74 mmol/mol).

### Predictive model and intervention

In an earlier Patient-Centered Outcomes Research Institute (PCORI)-funded study,^3^ our team at the Predictive Analytics and Comparative Effectiveness (PACE) Center at Tufts Medical Center and the American Medical Group Association (AMGA) developed an electronic health record (EHR)-based risk prediction model that provides individualized, point-of-care estimates to support targeted prevention for people with prediabetes. It calculates their risk of developing type 2 diabetes within three years and the benefit they would experience from the interventions evaluated in a landmark clinical trial, the DPP Study—offering either an intensive lifestyle program or metformin. We developed and validated the risk model using data from 2 million people with prediabetes in the Optum Labs Data Warehouse (OLDW), a de-identified database of healthcare claims and clinical data containing longitudinal health information on enrollees/patients, representing a diverse mixture of ages and geographic regions across the U.S. The model uses 11 variables that are typically available in an EHR (Table 1) and accommodates missing data. We used data from 3,081 people in the DPP Study to obtain risk-specific treatment effects.^3^

**Table 1.** EHR data elements included as predictors in type 2 diabetes risk model.

|  |
| --- |
| Age |
| Sex |
| Race (Black, Asian, White, other) |
| Smoking status (Current Former, Never) |
| Hypertension diagnosis (Yes, No) |
| Systolic blood pressure |
| Hemoglobin A1c |
| Fasting plasma glucose |
| Triglycerides |
| HDL cholesterol |
| Body mass index |

Note: Model design allows it to make predictions even when data for some variables are missing.

We integrated the risk prediction model into the clinical workflow, with automatic retrieval of model variables from PMA’s Allscripts EHR, using Galen eCalcs, which also offers other healthcare related calculators. This study focused on use of this model by primary care providers, at the point of care, to inform shared decision-making for people with prediabetes.

Using a modified RE-AIM framework ^7^, we evaluated the **<u>r</u>**each of the intervention, or the proportion of primary care patients in the system identified as having prediabetes for whom the tool was used. We evaluated **<u>e</u>**ffectiveness of the intervention, or knowledge and satisfaction with the risk calculator among both providers and patients, using pre/post surveys. We also measured **<u>a</u>**doption of the calculator by clinicians seeing eligible patients, and gauged **i**mplementation in terms of rates of referral to the DPP or prescribing metformin, stratified by the patient’s estimated risk of developing diabetes within 3 years (high, moderate, low). We measured **<u>m</u>**aintenance by collecting “reach” measures over time.

The implementation included all care teams across PMA’s seven primary care offices and provided communication about the need addressed by the model, training activities and resources, and other information and opportunities to provide input. Using evidence-based implementation science principles, we conducted patient and provider focus groups. The focus groups informed implementation, specifically how the patient- and provider-facing tools would look and be used in practice. Provider focus groups included seven to eleven primary care team members who would be using the risk calculator. Patient focus groups consisted of six to eight patients, drawn from organizational patient councils and patients referred by project team members. These sessions explored patients’ acceptance and understanding of the risk calculator and how best to communicate risk estimates.

As part of the implementation we conducted pre- and post-dissemination longitudinal cohort surveys with all providers across PMA’s primary care offices and with a random sample of patients. The patient sample was drawn from all patients with known prediabetes seen in clinic at PMA for well visits. Providers at PMA received pre- and post-dissemination surveys through email links to an online survey administered by SurveyMonkey. Patients received 12-item pre- and post-dissemination surveys through standard mail, with a study information sheet, a $2 bill, and an addressed, stamped envelope. Patient surveys were typically returned within 2-3 weeks of the primary care visit. Survey responses were stored and processed without linking to respondent identity information, to protect participants’ anonymity and confidentiality. All questionnaires are included in Supplements 1 and 2.

We used EHR data to assess the rates of risk calculator use by providers and to measure the rates of referrals to DPP programs and metformin prescribing. The study received IRB approval from Tufts Medical Center.

## Results

### Patient and Provider Focus Groups (pre-implementation)

Consistently, people with prediabetes said they wanted a personalized estimate of their risk of developing diabetes. Most focus group participants were able to quote the ages at which several family members developed type 2 diabetes. They were already thinking about their own personal risk in probabilistic terms, using the data they had available. Similarly, providers said they wanted individualized risk estimates for their patients, both to inform shared decision-making and to gain some sense of prioritization within the seemingly overwhelming number of people with prediabetes.

Given time constraints during the typical patient visit, providers expressed a preference to view the results of the predictive model within the routine patient encounter workflow in the EHR, rather than using a separate calculator. Patients emphasized the importance of information sharing and coordination among members of their care teams.

Without quantitative personalized risk estimates, providers were limited to qualitative discussion. Providers and patients alike perceived specific estimates of potential benefits of interventions as helpful for understanding risks and treatment options and motivating for behavior change.

### Pre- and Post-Surveys of Providers and Patients

Survey responses included 162 patients (24% response rate) and 24 providers (71% response rate) for pre-implementation and 171 patients (30% response rate) and 28 providers (82% response rate) for post-implementation surveys.

Surveys of providers and patients at PMA (Figure 1) showed that with the model, providers felt far more confident in their ability to estimate the risk of progression to diabetes for individual patients. Patients felt somewhat more confident in understanding their own risk. Without the model, patients’ confidence far exceeded that of providers. With the model, the providers experienced a much greater increase in confidence than did patients.

**Figure 1.**
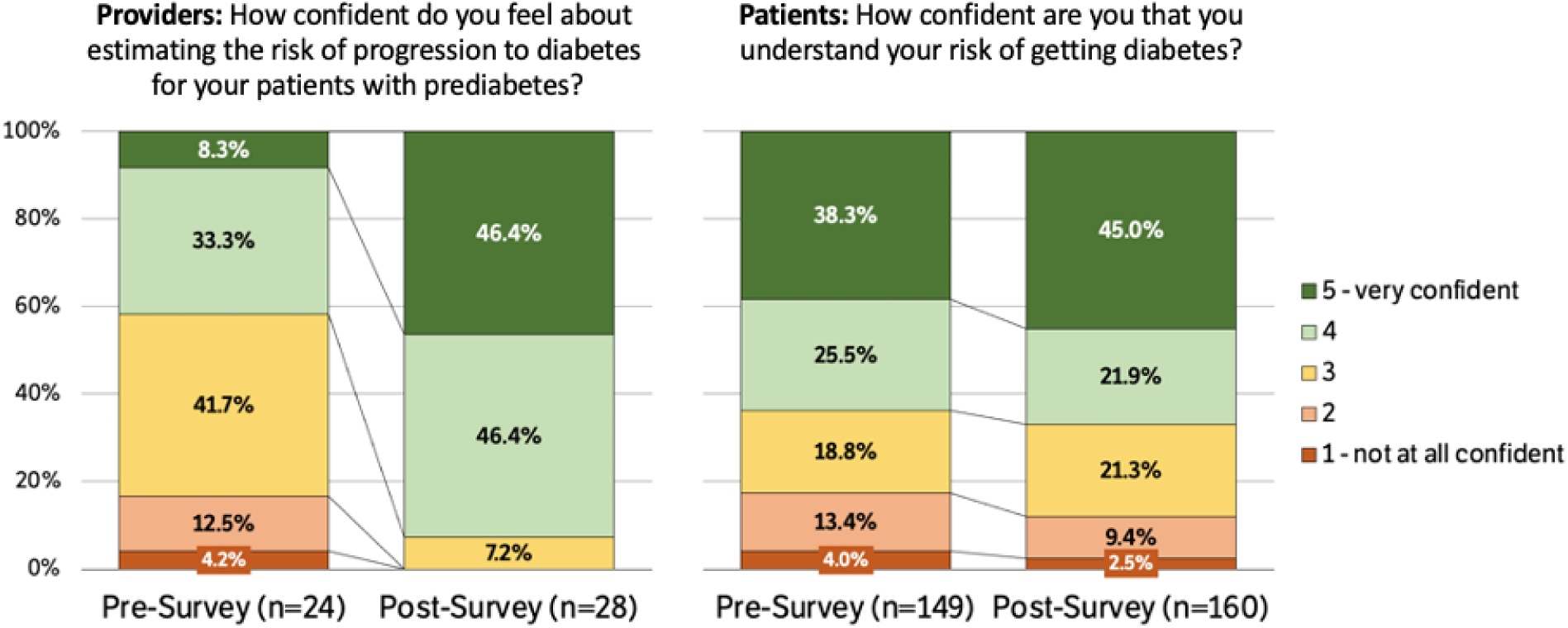
Pre- and post-survey responses of providers and patients at Premier Medical Associates: Confidence in estimating and understanding risk of progressing to type 2 diabetes

The model, when used by a clinician, helped patients understand their risks of developing type 2 diabetes (Figure 2). By far the patients’ most preferred source of information about their risk of diabetes was talking to their doctor (86% of patient respondents). Full tallies of survey responses are included in Supplements 3 and 4.

**Figure 2.**
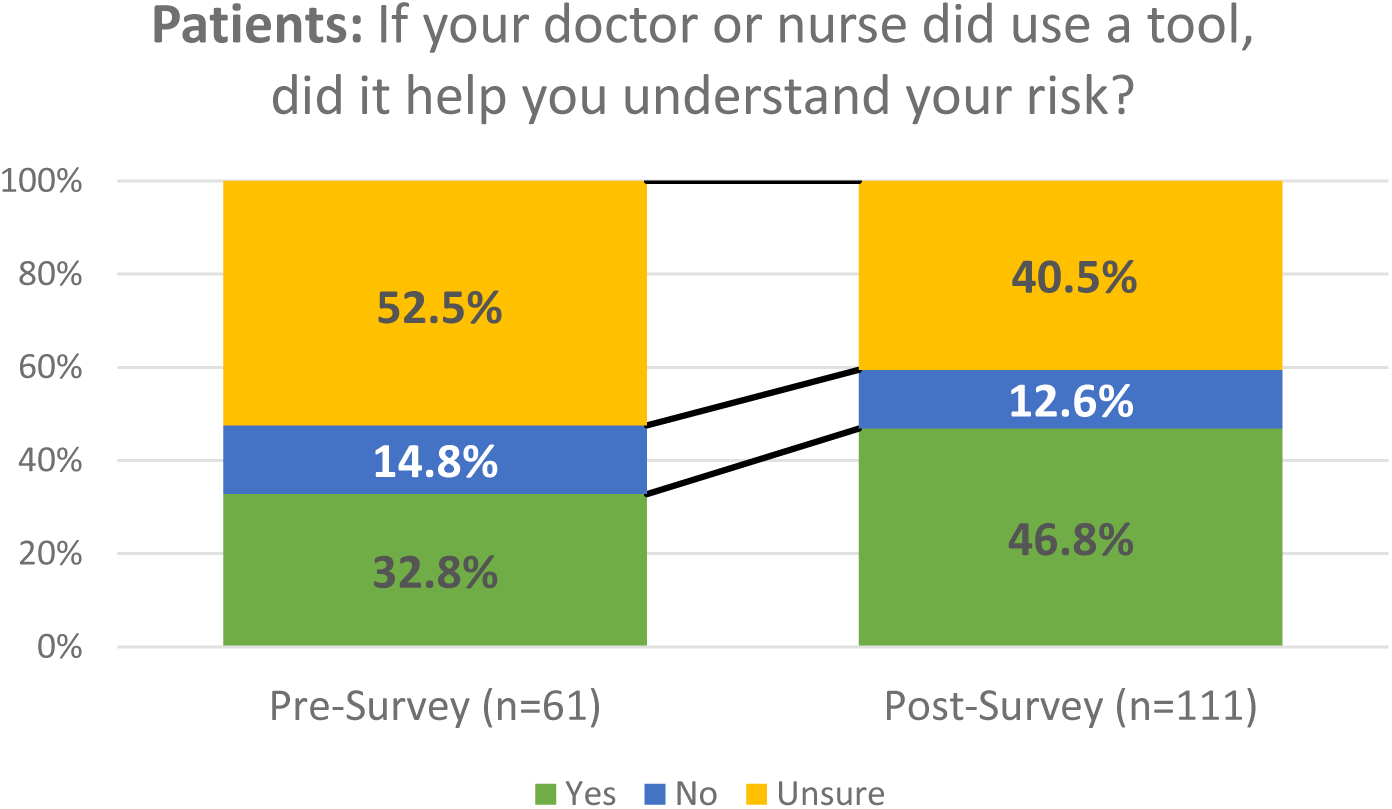
Pre- and post-survey responses of patients at Premier Medical Associates: Did tools for estimating risk of progression to type 2 diabetes used by your doctor or nurse help you understand your risk?

### Impact of Model Use by Providers

Between May 2018 and December 2020, 2,775 patients with prediabetes were seen in primary care at PMA. Of these patients, 78.9% had a calculation with the risk prediction tool completed, with rates increasing over time, from 52% in the initial 9 months to 79% over the full 32 months of the study (Table 2). Providers became increasingly aware of, and comfortable with, the model over time and reported finding it to be a useful tool for shared decision-making conversations with patients.

**Table 2.** Rates of risk calculator usage over time at Premier Medical Associates during the study period.

|  | 5/1/18–<br>1/31/19 | 5/1/18–<br>5/31/19 | 5/1/18–<br>8/31/19 | 5/1/18–<br>12/31/19 | 5/1/18–<br>12/13/20 |
| --- | --- | --- | --- | --- | --- |
| People with Prediabetes | 2,466 | 2,425 | 2,518 | 2,454 | 2,775 |
| Calculation Completed | 1,272 | 1,566 | 1,881 | 2,023 | 2,189 |
| Percent with Calculation | 52% | 65% | 75% | 82% | 79% |

Note: Patient counts reflect all patients with prediabetes, aged 18–75, with their last HbA1c during the time period in the range 5.7–6.4%, who had at least one visit with a primary care provider patients during the respective time period and were still active patients as of the end of the timeframe.

Before risk model implementation at PMA, no patient with prediabetes had been referred to the DPP program at the local YMCA, and fewer than 5% of people with prediabetes had been prescribed metformin. With the risk prediction model, preventive interventions increased greatly and became strongly risk stratified, with 62.3% of high-risk patients and 21.1% of moderate-risk patients receiving an intervention within 1 year (Table 3). Among high-risk patients receiving an intervention by 12/13/2020, 490 (71%) were referred to the DPP program and 202 (29%) were prescribed metformin.

**Table 3.** Risk-stratified rates of risk calculator usage, intervention and estimated type 2 diabetes risk at Premier Medical Associates.

| Risk level | Risk calculation completed |  | Intervention received |  | 3-year risk of type 2 diabetes |
| --- | --- | --- | --- | --- | --- |
|  | n | % | n | % |  |
| High | 1,109 | 50.7% | 690 | 62.3% | 41.6% |
| Moderate | 967 | 44.2% | 204 | 21.1% | 11.8% |
| Low | 113 | 5.2% | 10 | 9.7% | 3.8% |
| Total | 2,189 | 78.9%* | 904 | 41.3% | 19% |

\*N=2,775 patients with prediabetes
Data presented was collected during the time period 5/1/2018-12/13/2020. Intervention received was referral to DPP programs or metformin prescription.

Of 487 people with prediabetes who were referred to the DPP in the first few months after the model was implemented at PMA, 124 (25%) called the YMCA to inquire, and 64 (13%) enrolled in the DPP. During the year-long DPP program, these patients achieved an average weight loss of 7.4% (the “design goal” of the DPP is 7%).

## Discussion

This study demonstrated the successful implementation of a risk calculator in primary care. This tool calculated individualized risk of developing type 2 diabetes and the expected benefit of the DPP and metformin, based on 11 patient characteristics. Used in a shared decision-making process between patient and provider at the point of care, the calculator helped providers prioritize patients for prevention, facilitated communication, and successfully improved rates of engagement in their care among patients with prediabetes. A key component of the process was following implementation science principles to make the intervention design responsive to provider and patient needs and perspectives.

Nationally, very few people with prediabetes enroll in the DPP or receive metformin, although the results of the DPP Study were published more than 20 years ago.^8–10^ The high degree of heterogeneity of the risk of type 2 diabetes and of the benefits of diabetes prevention strategies presents a challenge to clinical decision-making, communication, care coordination, and ultimately engagement of patients in lifestyle change and adherence to interventions. Our study showed that personalized estimates of risk and benefit can engage patients and empower providers, informing shared decision-making around prevention choices. Providers and health systems need tools to help prioritize limited resources and increase patient treatment, referral, and adherence, through more targeted and tailored treatment recommendations. Because patients already value their providers as the best source of information, equipping providers with information to have these discussions enhances their effectiveness.

Even during the first year of the COVID-19 pandemic, the risk prediction model continued show great success at PMA. This has several important ramifications. First, health systems struggle to provide comprehensive care for all their patients and, with limited resources, need a way to prioritize patients at greatest need and identify those most likely to benefit from preventive interventions. This model stratifies patients by level of risk so that providers and health systems can use limited resources most efficiently. Second, even when prioritized, patients may be reluctant to engage in their own care, especially if they underestimate their risk of developing diabetes or view the DPP as labor-intensive or onerous. Third, many risk calculators are developed but sit on the shelf, gathering dust.

Our study findings from a single healthcare organization may not be fully generalizable across all regions and organizations. The impact of the risk model on clinical care depends on many factors such as local setting, populations, and implementation choices. Future studies should extend evaluation to implementations in other health care systems and regions.

## Conclusions

This study demonstrates the potential impact of successful implementation of a validated risk model in routine clinical practice. Work is underway to develop technology using the Fast Healthcare Interoperability Resources (FHIR) standard to integrate a cloud-hosted version of the calculator into Epic and other leading EHRs, facilitating wider dissemination and improved care coordination. Future directions for research include implementation at Mercy, a 2,900-provider integrated system serving patients across four states (AR, KS, MO, OK), and other large health systems. This demonstration of translating research into practice provides a path for personalized medicine to become a reality for patients.

## Data Availability

All data generated during and/or analyzed during the current study are available from the corresponding author upon reasonable request.

## Declarations

### Ethics approval and consent to participate

The project was approved by the Tufts Health Sciences Institutional Review Board. Informed consent was obtained from participants.

### Consent for publication

Not applicable.

### Availability of data and material

The datasets generated during and/or analyzed during the current study are available from the corresponding author upon reasonable request.

### Competing interests

All authors declare no conflicts of interest relevant to this article.

### Funding

Research reported in this publication was funded through a Patient-Centered Outcomes Research Institute (PCORI) Award (DI-1604-35234). The statements in this work are solely the responsibility of the authors and do not necessarily represent the views of the Patient-Centered Outcomes Research Institute (PCORI), its Board of Governors or Methodology Committee. Dr. Kent was funded by a National Institutes of Health (NIH)/National Center for Advancing Translational Sciences (NCATS) grant (UM1TR004398-01).

### Authors’ contributions

NO, JC, EC, and DMK drafted the manuscript. EC, JC, and DMK oversaw and designed the study. JC and DMK obtained funding for the research. FC and CK contributed to discussion, and reviewed and edited the manuscript. All authors contributed to data access and collection, analysis, development of conclusions, reviewed and contributed significantly to the final manuscript. All authors approve the final version of the manuscript. DMK is the guarantor of this work and, as such, had full access to all the data in the study and takes responsibility for the integrity of the data and the accuracy of the data analysis.

## Acknowledgements

Not applicable.

## Notes

### Competing Interest Statement

The authors have declared no competing interest.

### Funding Statement

This study was funded through a Patient-Centered Outcomes Research Institute (PCORI) Award (DI-1604-35234). The statements in this work are solely the responsibility of the authors and do not necessarily represent the views of the Patient-Centered Outcomes Research Institute (PCORI), its Board of Governors or Methodology Committee. Dr. Kent was funded by a National Institutes of Health (NIH)/National Center for Advancing Translational Sciences (NCATS) grant (UM1TR004398-01).

### Author Declarations

The Institutional Review Board of Tufts Health Sciences gave ethical approval for this work.

### Summary of Updates

Funding Statement updated to include NIH funding of Dr. David Kent.

